# Association of Sociodemographics with Out-of-Pocket Expenditure and Underinsurance in Skin Cancer Survivors

**DOI:** 10.1101/2021.07.17.21251600

**Authors:** Mohammad A. Karim, Robert L. Ohsfeldt, Nima Khodakarami, Hye-Chung Kum

## Abstract

**Background:** The sociodemographic predictors of either out-of-pocket expenditure (OOP) or underinsurance among skin cancer survivors is not well reported in the literature. In this study we estimated all-cause healthcare related OOP expenditure and probability of underinsurance among insured skin cancer survivors and identified the sociodemographic predictors of these measures.

**Data and Method:** We pooled Medical Expenditure Panel Survey (MEPS) data from 2011 to 2015 and identified skin cancer using Clinical Classifications Software (CCS) code. Only adult (≥18 years) skin cancer survivors with full year insurance coverage were included in our study (n = 1825). We used a generalized linear model (GLM) with log link and gamma distribution to estimate OOP and a logit model to estimate the probability of underinsurance. We estimated the Average Marginal Effect (AME) to quantify the variations and their statistical significance between reference level and other levels of each predictor. Our analyses accounted for the complex survey design of MEPS.

**Results:** The average all-cause OOP was $1766 per person per year for a skin cancer survivor. Among all skin cancer survivors, females, those aged 60-64 years, with some college education or a degree, with income ≥400% of federal poverty level (FPL) and with non-managed-care private insurance incurred significantly higher OOP expenditure compared to their respective counterparts. In terms of underinsurance, females and those aged 60-64 years had higher probability, whereas, survivors with non-white race/ethnicity and income 200% of FPL or higher had lower probability of being underinsured compared to their respective counterparts.

**Conclusion:** Our study demonstrates that OOP expenditure and underinsurance varies significantly by sociodemographic factors among skin cancer survivors.

## 1.1. INTRODUCTION

Skin cancer is the most common cancer in the United States, with an estimated 3.3 million new patients diagnosed with non-melanoma skin cancer (NMSC) and an estimated 90 thousand new patients diagnosed with melanoma in 2018 (US Preventive Services Task Force et al., 2018). Although mortality from certain skin cancers, like NMSC, is lower than other cancer types, skin cancer is responsible for more than 60% of total mortality caused by all skin disorders (Lim et al., 2017). Melanoma caused more than 6800 estimated deaths in 2020 and NMSC causes around 2000 deaths per year in the US (American Cancer Society, 2020a, 2020b). One in five Caucasian American is expected to be diagnosed with NMSC by the time they reach the age of 70 (Stern, 2010). The aggregated financial burden of skin cancer is also substantial, with a staggering total national burden of $8.1 billion per year incurred during the 2007-2011 period (Guy, Machlin, Ekwueme, & Yabroff, 2015).

Several studies have estimated the cost of skin cancer using Medicare data (Ruiz, Morgan, Zigler, Besaw, & Schmults, 2019; Seidler, Pennie, Veledar, Culler, & Chen, 2010) and national survey data (J. G. Chen et al., 2001). These studies, however, are not reflective of the total financial burden experienced by skin cancer survivors, because, they usually focus only on cancer related healthcare costs rather than all-cause healthcare costs. Moreover, out-of-pocket expenditure (OOP) is sparsely reported in skin cancer cost related literature. Adequacy of insurance coverage is another under-investigated topic in context of skin cancer. To the best of our knowledge, no reported study to-date has investigated the sociodemographic predictors of either OOP expenditure or underinsurance among skin cancer survivors. Among the recent studies, Chen et al. reported expenditure variations related to treatment practice differences among NMSC survivors with Medicare coverage (J. T. Chen, Kempton, & Rao, 2016), whereas, Ruiz et al. reported results for a similar analysis among Medicare population for all skin cancer subtypes (Ruiz et al., 2019). However, neither of these studies investigated the sociodemographic variation in OOP expenditure among skin cancer survivors.

In the current study, we estimated all-cause healthcare related OOP expenditure and probability of underinsurance among insured skin cancer survivors and identified the sociodemographic predictors of these measures.

## 1.2. METHODS

### 1.2.1. Data Source

In the current study, Medical Expenditure Panel Survey (MEPS) data was used (years: 2011 to 2015). MEPS provides several publicly available datasets, among which the Full Year Consolidated (FYC) file (Agency for Healthcare Research and Quality, 2017b) and the Medical Conditions (MC) file were used for our analysis (Agency for Healthcare Research and Quality, 2017a). The FYC file for a given year provides sociodemographic and expenditure data, and the MC file provides medical condition related data for the survey sample for that year (Agency for Healthcare Research and Quality, 2017a, 2017b). In our analysis, MC file for each year was linked to the FYC file using the common individual level identifier and all health-related expenditures incurred by a skin cancer survivor were aggregated. This was repeated for each year from 2011 to 2015 and the resultant year-specific linked files were pooled together to generate the final analytic file.

### 1.2.2. Study Sample

Skin cancer cases were identified using Clinical Classifications Software (CCS) codes 22 (melanoma) and 23 (NMSC) from the MEPS MC file. While pooling linked FYC-MC data for the years 2011 to 2015, person level probability weights were adjusted by dividing the weights by the number of pooled years, as recommended by MEPS (Agency for Healthcare Research and Quality, 2017c). Since the probability of underinsurance warrants inclusion of the individuals with full year insurance coverage, only those individuals were included. All included individuals were adult skin cancer survivors aged 18 years or older.

### 1.2.3. Measures

#### Expenditure and income

MEPS-reported out-of-pocket expenditures for all health services incurred by skin cancer survivors were summed to obtain the total all-cause OOP expenditure. To determine underinsurance, MEPS-reported family income was used. Consumer Price Index (CPI) was used to inflate all dollar values to 2018 US$.

#### Underinsurance

In accordance with the prior literature, the underinsurance indicator variable was defined based on the ratio of total OOP expenditure to family income. An individual with family income <200% of federal poverty level (FPL) was deemed underinsured if their total OOP was ≥5% of the family income, and an individual with family income ≥200% of FPL was deemed underinsured if their total OOP was ≥ 10% of family income (Magge, Cabral, Kazis, & Sommers, 2013; Schoen, Doty, Robertson, & Collins, 2011).

#### Predictors

Sociodemographic variables age, sex, race/ethnicity, marital status, income level, education and insurance status were the predictor variables. The number of MEPS priority conditions were coded as a categorical variable. To account for potential confounding effect of health status variation and regional variation, self-reported health status and census region variables were added in regression models. All independent variables in each model were categorical (**Table 1**). Because skin cancer survivors are overwhelmingly white, the race/ethnicity variable had only two categories (non-Hispanic white [ref.], non-white); any individual with race/ethnicity other than non-Hispanic white were grouped under the ‘non-white’ category.

**Table 1.** Skin cancer survivors by sociodemographic factors.

| Variable | Categories | all skin cancer<br>(unweighted<br>n=1825) | NMSC<br>(unweighted<br>n=1566) | Melanoma<br>(unweighted<br>n=259) | *p*<sup>†</sup> |
| --- | --- | --- | --- | --- | --- |
| age | 18-49 years | 6.9% | 6.3% | 10.3% | 0.048 |
|  | 50-59 years | 16.0% | 16.0% | 16.5% |  |
|  | 60-64 years | 11.7% | 11.1% | 15.7% |  |
|  | 65-74 years | 31.2% | 31.0% | 32.8% |  |
|  | 75-84 years | 24.4% | 25.1% | 20.1% |  |
|  | ≥85 years | 9.7% | 10.5% | 4.6% |  |
| sex | Male | 52.0% | 52.2% | 50.9% | 0.785 |
|  | Female | 48.0% | 47.8% | 49.1% |  |
| race/ethnicity | non-Hispanic white | 96.8% | 97.8% | 90.1% | 0.000 |
|  | non-white | 3.2% | 2.2% | 9.9% |  |
| marital status | Not Married | 32.6% | 32.0% | 35.9% | 0.377 |
|  | Married | 67.4% | 68.0% | 64.1% |  |
| education | HS education/diploma | 31.2% | 32.5% | 23.4% | 0.057 |
|  | Some college | 26.7% | 25.7% | 32.9% |  |
|  | College degree or above | 42.1% | 41.8% | 43.8% |  |
| income level | <200% FPL | 20.3% | 20.8% | 17.4% | 0.431 |
|  | 200%-400% FPL | 23.0% | 23.1% | 21.8% |  |
|  | ≥400% FPL | 56.7% | 56.1% | 60.8% |  |
| insurance status | Private – MC | 15.3% | 14.9% | 17.9% | 0.545 |
|  | Private - non MC | 61.3% | 61.3% | 61.4% |  |
|  | Public | 23.3% | 23.8% | 20.7% |  |
| number of comorbidities | none | 6.5% | 5.7% | 11.3% | 0.059 |
|  | one | 11.8% | 11.4% | 14.0% |  |
|  | two | 17.0% | 17.2% | 16.1% |  |
|  | three or more | 64.7% | 65.7% | 58.7% |  |
| census region | northeast | 16.3% | 16.4% | 15.6% | 0.797 |
|  | midwest | 24.2% | 24.1% | 25.1% |  |
|  | south | 36.6% | 37.2% | 33.2% |  |
|  | west | 22.8% | 22.3% | 26.1% |  |
| health status | poor or fair | 14.9% | 14.4% | 17.7% | 0.445 |
|  | good | 28.4% | 28.3% | 29.1% |  |
|  | very-good or excellent | 56.7% | 57.3% | 53.3% |  |

Abbreviations: HS, high school; MC, Managed care.
Note: Survey weighted descriptive statistics based on the analysis of Medical Expenditure Panel Survey data (2011-2015).
<sup>†</sup>*p*-value from chi-squared test of independence (survey-weighted) among categorical variables across NMSC and melanoma.

### 1.2.4. Analysis

#### Statistical analysis

Descriptive analysis was conducted to identify sample characteristics. Mean unadjusted OOP expenditure for all survivors and survivors with a positive expenditure were estimated applying survey weights. Additionally, covariate adjusted analyses were performed using generalized linear model (GLM) with log link and gamma distribution, incorporating all key independent variables, along with the potential confounding variables. The GLM model was used to assess the simultaneous effect of the independent variables and to quantify the covariate adjusted mean OOP (Deb & Norton, 2018). Probability of underinsurance was estimated using a logistic models using the same set of covariates. The significance level was set at 5%. Incremental effect size across categories of a predictor and their statistical significance were determined using Average Marginal Effect (AME) for both GLM and logistic models (Mize, 2019). All analyses in our study were performed accounting for the complex survey design of MEPS (Agency for Healthcare Research and Quality, 2017b; Machlin, Yu, & Zodet, 2005), using Stata13.1 software (StataCorp, College Station, TX).

## 1.3. RESULTS

### 1.3.1. Descriptive Statistics

Our sample included 1825 skin cancer survivors among which 1566 were NMSC survivors and 259 were melanoma survivors. There were significant differences in age and race/ethnicity between NMSC and melanoma cases. Notably, proportion of survivors aged 18-49 years was higher, whereas the proportion of survivors aged ≥85 years was lower in the melanoma cohort. With respect to race/ethnicity, percentage of non-Hispanic whites was lower in the melanoma cohort **(Table 1)**.

In unadjusted analysis, when only non-zero cases were considered, survey weighted all-cause OOP in 2018$ were higher for melanoma ($1808, SE $184) vs. NMSC ($1754, SE $117). OOP for only NMSC cases was slightly lower than all skin cancer cases combined ($1761, SE $106). When both zero and non-zero expenditure cases were considered together, similar dollar values for each subtype was observed (**Figure 1**).

**Figure 1:**
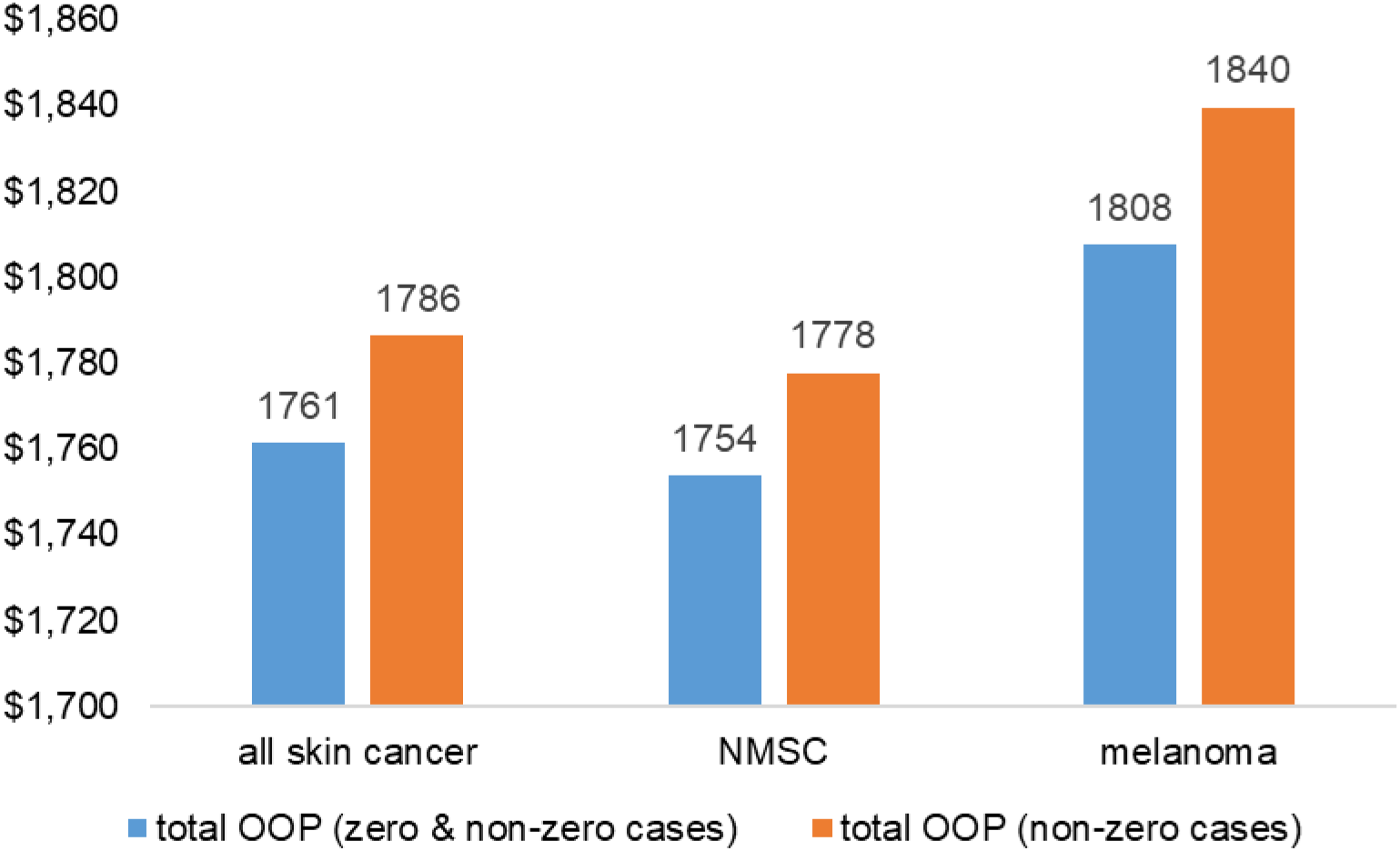
Unadjusted mean all-cause total OOP incurred by skin cancer survivors, 2011-2015 (2018$)

### 1.3.2. Adjusted OOP Expenditure and Underinsurance

Average adjusted predictions (AAP) from the analysis showed that among all skin cancer survivors, average all-cause OOP expenditure was $1766 per person per year, and the mean probability of underinsurance was 13.2%. The average adjusted all-cause OOP value and probability of underinsurance remained very similar with values of $1763 and 13.2%, respectively, when only the NMSC cases were considered. Covariate adjusted OOP and underinsurance analysis were not conducted for melanoma skin cancer cases separately due to the small sample size.

### 1.3.3. Sociodemographic Factors Associated With OOP Expenditure

Among all skin cancer cases, survivors aged 60-64 years spent $623 more than survivors aged 18-49 years [reference] (p<0.05); females spent $390 more than males [ref.] (p<0.01); survivors with some college education and with a college degree spent $374 and $910 more than survivors with no college education [ref.] (both p<0.01), respectively; survivors with income ≥400% of FPL spent $435 more than those with income <200% of FPL [ref.] (p<0.05); and, survivors with non-managed-care private insurance spent $334 more than those with managed-care private insurance [ref.] (p<0.05), on-average per person per year (**Table 2**).

**Table 2.**
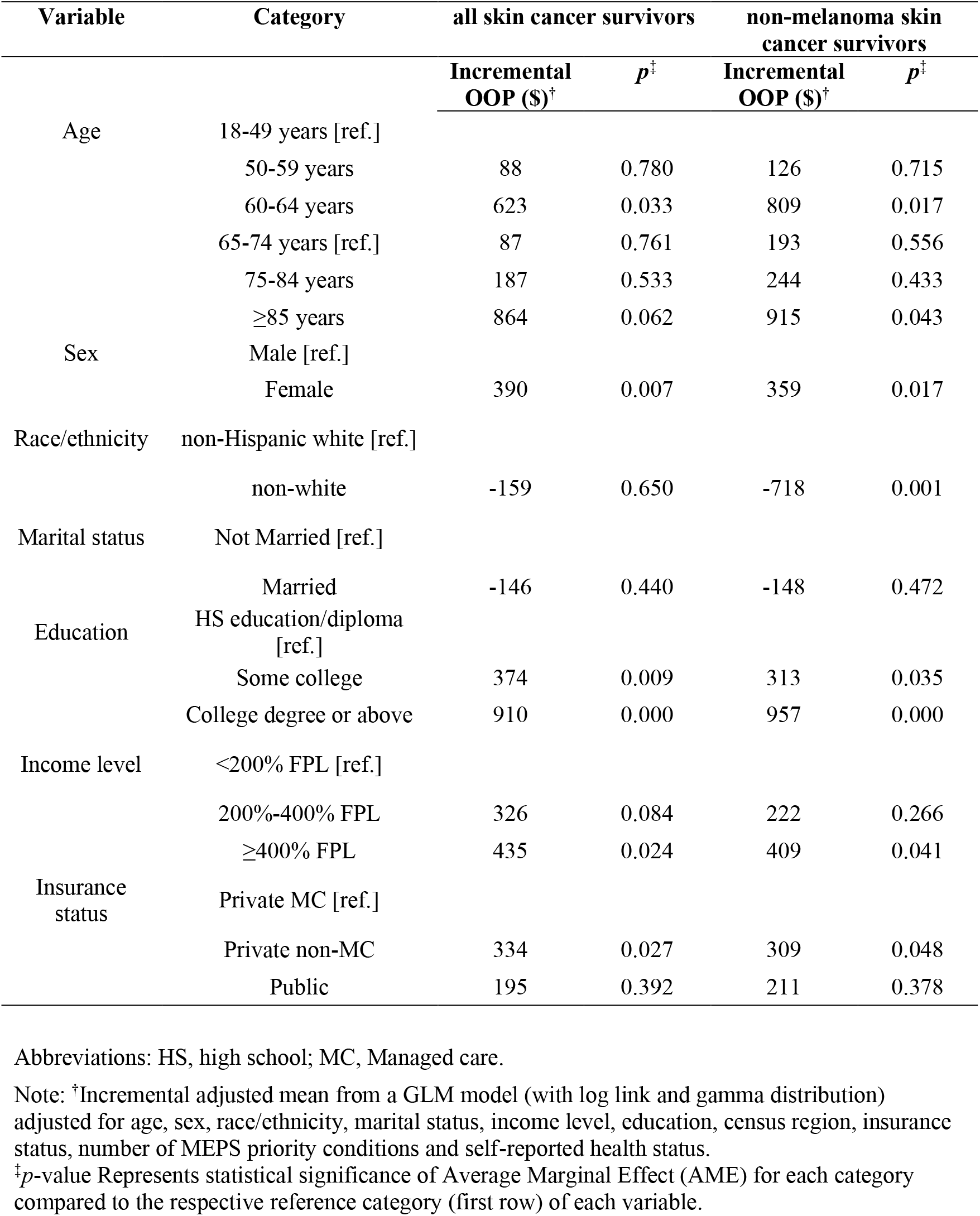
Variation in all-cause total OOP by sociodemographic factors among skin cancer survivors (weighted).

When only NMSC cases were considered, the same sociodemographic groups with significantly different OOP in all-skin cancer analysis showed similar variation vs their respective counterparts; with the only exception of non-white race/ethnicity. Although all-cause OOP did not vary significantly among non-Hispanic whites vs non-whites in the all skin cancer analysis, significantly lower OOP were observed for non-whites when only NMSC cases were considered **(Table 2)**.

### 1.3.4. Sociodemographic Factors Associated With Underinsurance

Among all skin cancer cases, females had 3.9 percentage point (pp) higher probability of underinsurance vs males [ref.] (p<0.01); survivors aged 60-64 years had 8.5 pp higher probability vs survivors aged 18-49 years [ref.] (p<0.05); those of non-white race/ethnicity had

8.3 pp lower probability vs non-Hispanic whites [ref.] (p<0.01), and, those with income 200%-400% of FPL and ≥400% of FPL had 38.3 pp and 47.6 pp lower probability (both p<0.01), respectively, vs those with income <200% of FPL [ref.] of being underinsured.

When only NMSC cases were considered, the same sociodemographic factors, namely age between 60 and 64 years, female sex, non-white race/ethnicity, income 200%-400% of FPL and ≥400% of FPL were associated with variations in underinsurance in the same directions **(Table 3)**.

**Table 3.** Variation in probability of underinsurance by sociodemographic factors among skin cancer survivors (weighted).

| Variable | Category | all skin cancer survivors |  | non-melanoma skin cancer survivors |  |
| --- | --- | --- | --- | --- | --- |
|  |  | Incremental probability of underinsurance (%)<sup>†</sup> | *p*<sup>‡</sup> | Incremental probability of underinsurance (%)<sup>†</sup> | *p*<sup>‡</sup> |
| Age | 18-49 years [ref.] |  |  |  |  |
|  | 50-59 years | 1.5% | 0.671 | 2.7% | 0.390 |
|  | 60-64 years | 8.5% | 0.029 | 11.9% | 0.001 |
|  | 65-74 years [ref.] | 2.4% | 0.445 | 4.3% | 0.129 |
|  | 75-84 years | 2.0% | 0.517 | 4.0% | 0.147 |
|  | ≥85 years | 5.4% | 0.154 | 6.6% | 0.056 |
| Sex | Male [ref.] |  |  |  |  |
|  | Female | 3.9% | 0.003 | 3.3% | 0.011 |
| Race/ethnicity | non-Hispanic white [ref.] |  |  |  |  |
|  | non-white | -8.3% | 0.000 | -10.9% | 0.000 |
| Marital status | Not Married [ref.] |  |  |  |  |
|  | Married | 0.0% | 0.979 | 0.6% | 0.686 |
| Education | HS education/diploma [ref.] |  |  |  |  |
|  | Some college | 2.7% | 0.073 | 2.6% | 0.073 |
|  | College degree or above | 2.6% | 0.177 | 2.1% | 0.352 |
| Income level | <200% FPL [ref.] |  |  |  |  |
|  | 200%-400% FPL | -38.3% | 0.000 | -39.6% | 0.000 |
|  | ≥400% FPL | -47.6% | 0.000 | -48.1% | 0.000 |
| Insurance status | Private MC [ref.] |  |  |  |  |
|  | Private non-MC | 1.3% | 0.526 | 1.3% | 0.572 |
|  | Public | -0.2% | 0.911 | 0.3% | 0.884 |

Abbreviations: HS, high school; MC, Managed care.
Note: <sup>†</sup>Incremental adjusted mean from a logistic model; adjusted for age, sex, race/ethnicity, marital status, income level, education, census region, insurance status, number of MEPS priority conditions and self-reported health status.
<sup>‡</sup>*p*-value Represents statistical significance of Average Marginal Effect (AME) for each category compared to the respective reference category (first row) of each variable.

## 1.4. DISCUSSION

In the current study, an all-cause OOP expenditure approach was adopted to investigate the financial burden on insured skin cancer survivors. Our analyses showed that around 13% of the skin cancer survivors experienced underinsurance and average all-cause OOP was $1766 per person per year.

A prior study by Ruiz et al. using 2013 Medicare Limited Dataset Standard Analytic File 5% sample estimated average cancer attributable cost of melanoma, basal cell carcinoma (BCC) NMSC and squamous cell carcinoma (SCC) NMSC per person per year to be $1241, $689 and $649 respectively (Ruiz et al., 2019). In our analysis, OOP cost estimates for melanoma and NMSC are higher than the total cost estimates reported by Ruiz et al (Ruiz et al., 2019). This is probably because we did not differentiate between cancer attributable and non-attributable costs.

Another study by Guy et al. reported higher cancer attributable average per person cost for melanoma of $4780 using 2007-2011 MEPS data (Guy et al., 2015). In their study, Guy et al. also reported the changing trend in NMSC and melanoma prevalence among males and females across age groups over time; however, they did not report cost across demographics or socioeconomic status (Guy et al., 2015). In the current study we observed sociodemographic variation in OOP expenditure among insured skin cancer survivors, with females and college educated individuals spending significantly higher OOP amounts. This indicates that these insured subgroups of survivors may need special attention in terms of financial assistance. Additionally, we observed significantly higher OOP expenditure among survivors with income ≥400% of FPL.

There is very limited evidence in the literature concerning variation in either total or OOP expenditure across insurance types among skin cancer survivors. Guy et al. reported that 43.4% and 41.1% of total national cost burden on skin cancer is borne by private insurance and Medicare respectively (Guy et al., 2015); however, the association between insurance types and OOP expenditure was not reported. Current study found that OOP expenditure varied significantly with insurance type and skin cancer survivors with private non-managed care insurance incurred higher OOP expenditure compared to those with private managed care insurance. This was observed among all skin-cancer cases, as well as, among only NMSC cases. This may indicate that private managed care plans may contribute in curbing OOP costs among skin cancer survivors. The role of managed care plans in containing OOP burden should be explored further in future studies.

Of note, our study identified sociodemographic variation in underinsurance among skin cancer survivors. Consistent with the higher incurred OOP, the probability of underinsurance was higher among survivors aged 60-64 years and females. Despite their higher OOP expenditure, survivors with income 200% of FPL and higher had significantly lower probability of underinsurance. Additionally, non-white survivors had lower probability of underinsurance.

There are certain limitations in the current study. Since MEPS is a survey-based dataset, recall bias may be present in the reported data (Althubaiti, 2016). Additionally, clinical stage or cancer phase could not be accounted for in the study due to the lack of reported clinical information in MEPS (Agency for Healthcare Research and Quality, 2017b). Moreover, the cost analysis in this study focused on all health-related costs and does not represent cancer-attributable costs. Our study was cross sectional in nature, and as such, before- and after-ACA change in cost trend was not identified.

Although certain skin cancer types may not be as deadly as other cancers, patients diagnosed with skin cancer may be subject to significant out-of-pocket burden. In this study we found that more than 13% of all insured skin cancer survivors experience underinsurance. Sociodemographic variations in OOP expenditure and underinsurance in this study highlights the varying financial challenges faced by different sub-groups of skin cancer survivors. Our findings underscore the necessity of needs-specific interventions to contain financial burden on skin cancer survivors.

## Data Availability

Data used in this study is available from Agency for Healthcare Research and Quality (AHRQ) - Medical Expenditure Panel Survey (MEPS) website.

https://meps.ahrq.gov/data_stats/download_data_files.jsp

